# Minimal in-school SARS-CoV-2 transmission with strict mitigation protocols at two independent schools in Nashville, TN

**DOI:** 10.1101/2021.11.09.21266140

**Authors:** Lauren S. Peetluk, Peter F. Rebeiro, Kathryn M. Edwards, Ritu Banerjee, Simon A. Mallal, David M. Aronoff, Loren Lipworth, Sophie E. Katz

**Affiliations:** Department of Medicine, Division of Infectious Diseases, Vanderbilt University Medical Center, Nashville, TN; Department of Medicine, Division of Epidemiology, Vanderbilt University Medical Center, Nashville, TN; Department of Biostatistics, Vanderbilt University Medical Center, Nashville, TN; Department of Pediatrics, Division of Infectious Diseases, Vanderbilt University Medical Center, Nashville, TN

**Author notes:** **Corresponding author:** Lauren Peetluk, PhD, MPH: A2209 Medical Center North, 1161 21st Ave S, Nashville, TN 37203. **Alternate corresponding author:** Sophie Katz, MD, MPH: D-7235 Medical Center North, 1161 21st Ave S, Nashville TN 37203.

**Keywords:** COVID-19, schools, transmission, disease outbreaks, attack rate

## Abstract

**BACKGROUND:** The COVID-19 pandemic has greatly impacted school operations. To better understand the role of schools in COVID-19 transmission, we evaluated infections at two independent schools in Nashville, TN during the 2020-2021 school year.

**METHODS:** The cumulative incidence of COVID-19 within each school, age group, and exposure setting were estimated and compared to local incidence. Primary attack rates were estimated among students quarantined for in-school close contact.

**RESULTS:** Among 1401 students who attended school during the study period, 98 cases of COVID-19 were reported, corresponding to cumulative incidence of 7.0% (95% confidence interval (CI): 5.7-8.5). Most cases were linked to household (58%) or community (31%) transmission, with few linked to in-school transmission (11%). Overall, 619 students were quarantined, corresponding to >5000 person-days of missed school, among whom only 5 tested positive for SARS-CoV-2 during quarantine (primary attack rate: 0.8%, 95% CI: 0.3, 1.9). Weekly case rates at school were not correlated with community transmission.

**CONCLUSION:** These results suggest that transmission of COVID-19 in schools is minimal when strict mitigation measures are used, even during periods of extensive community transmission. Strict quarantine of contacts may lead to unnecessary missed school days with minimal benefit to in-school transmission.

## BACKGROUND

The coronavirus disease 2019 (COVID-19) pandemic has had a major impact on K-12 school operations, with active debate surrounding whether schools represented a source of transmission to children and their teachers. Many schools closed and shifted to virtual learning, while others remained open with mitigation strategies in place. Existing evidence shows that children are less likely to become symptomatic, be hospitalized, or die from severe acute respiratory syndrome coronavirus 2 (SARS-CoV-2) than adults.(1) Prior to widespread vaccination in the United States, only 10% of all COVID-19 cases and 0.1% of all COVID-19 deaths reported were among children and adolescents aged 5 through 17 years, (2) an age group that makes up 15% of the population.(3) As vaccination rates increase among adults, children may account for larger proportions of cumulative cases and deaths.

Some reviews and well-designed studies find minimal evidence that in-person learning or school re-openings drive community transmission of COVID-19, especially when strict mitigation measures are in place.(4–7) However, other studies report that transmission may be more common among children in school environments than community settings, and transmission risk among school children increases with older age groups.(8,9) Other reports suggest that when transmission or outbreaks in schools did occur, they were often driven by teachers or staff, rather than student-to-student or student-to-staff transmission.(1,10) Overall, there remains uncertainty about how to safely and effectively operate schools with in-person learning, especially with regard to implementing mitigation measures. In this report, we describe the distribution and characteristics of COVID-19 cases at two independent schools in Nashville, Tennessee that remained open throughout the 2020-2021 school year with strict mitigation protocols in place.

## METHODS

Full details on data collection, mitigation strategies, contact tracing, quarantine protocols, and study definitions are included in **Supplementary File 1**.

### Participants and data sources

#### Data collection

Data were collected from parents via phone or email interview by the school health team at two independent schools in Nashville, TN (Davidson County). School A included students in Kindergarten through 12^th^ grade and School B included students in pre-Kindergarten through 8^th^ grade. However, for School B, only Kindergarten through 8^th^ grade were included in the study to increase comparability with Davidson County data collection (starting at age 5), whereas School A contributed data for Kindergarten through Grade 12. Data were collected from the beginning of the school year through February 12, 2021 and were retrospectively reviewed for this study. School A staggered their in-person opening by grade level, starting on September 9 with Kindergarten and concluding on October 12, 2020 with high school seniors returning. The in-person school year at School B began for all students on August 18, 2020. During these periods, all suspected and known cases of COVID-19 among students and faculty/staff were reported to the school health team. One member of the COVID-19 Medical Advisory Board from each school (epidemiologist for School A, infectious disease physician for School B), adjudicated the reports. A confirmed case was defined as having a positive polymerase chain reaction (PCR) test for SARS-CoV-2 infection.

### Procedure

#### Mitigation strategies

Each school retained in-person learning with strict mitigation measures throughout the study period. Briefly, these mitigation measures included daily symptom and exposure screening for all students, faculty and staff; physical distancing (desks spaced 3-6 feet apart with plexiglass); outdoor lunch and recess (weather permitting, or indoor silent lunch); strict hand hygiene; universal masking of all ages at all times; cohorting of classes with restricted sizes and with minimal mixing between cohorts (with the exception of School A grades 11-12; and restricted visitor access with COVID-19 symptom and exposure screening.

Families were additionally asked to keep students at home starting from the first sign or symptom of COVID-19 in any household member or suspected contact while PCR test result was pending. Sports teams practiced at School A with strict mitigation measures, informed by the American Academy of Pediatric guidelines, but were suspended at School B.(11)

#### Contact tracing and quarantine procedures

Individuals who attended school during the 48 hours prior to symptom onset or positive PCR test confirming SARS-CoV-2 infection were considered infectious while at school, and their in-school close contacts (≥15 minutes within 6 feet) were quarantined according to school policy. When it was not possible to discern the exact distance between students or contact patterns, such as among lower school students, whole classrooms were quarantined. Additionally, students with close contact to a confirmed COVID-19 case at home or in the community were quarantined.

At School A, the quarantine period was 14 days from positive test or onset of symptoms through December 5, 2020, when CDC issued revised quarantine guidelines. After that, the period of quarantine was reduced to 10 days. At School B, the quarantine period was 14 days through December 5, 2020. After that, the quarantine period changed to 10 days for all close contacts, with the option to reduce to 7 days if the individual remained symptom free and had a negative SARS-CoV-2 PCR test after day 5. The families of students under quarantine were asked to report any positive SARS-CoV-2 test results to the school.

#### Local COVID-19 data

Davidson County data were obtained from the Tennessee Department of Health Website (https://www.tn.gov/health/cedep/ncov/data/downloadable-datasets.html), including number of new cases per day for the whole population and among the subset of school-aged children and adolescents aged 5-18 years. Daily incidence rates per 100,000 population were calculated based on 2019 Census data, which estimated 694,150 people in the county, 16.1% of whom were between the ages of 5 and 18 years.(12)

### Definitions

Students were grouped according to three grade level groups: lower school (Kindergarten through 4^th^ grades), middle school (5^th^ through 8^th^ grades), and high school (9^th^ through 12^th^ grades). The exposure setting of the cases reported to each school was defined as in-school (epidemiologic link to a confirmed COVID-19 case who attended school during their infectious period), home (epidemiologic link to a confirmed COVID-19 case at home), or community (no known epidemiologic link to in-school or home transmission with or without the known source of community transmission). Infectious period was defined as the 48 hours prior to symptom onset or positive PCR test for SARS-CoV-2 infection. Cases were additionally considered as symptomatic or asymptomatic based on whether they developed COVID-19 related symptoms at any point during their illness. A case cluster was defined as at least two epidemiologically linked cases who were diagnosed within 14 days of each other.

### Data analysis

Characteristics of COVID-19 cases, including exposure setting, symptom status, and number quarantined (if infectious while at school) were described and compared between grade groups. Comparisons were evaluated with Chi-square tests for categorical variables, Fisher’s exact test for categorical variables with excepted cell counts were <5, and Kruskal-Wallis test for continuous variables.

The cumulative incidence of COVID-19 during the study period was estimated as the total number of positive COVID-19 tests reported to both schools combined divided by total number of students attending schools in-person during the study period. We additionally evaluated cumulative incidence by school, grade level groups, and exposure setting. The school-based cumulative incidence calculations were descriptively compared to the cumulative incidence among school-aged children in Davidson County, estimated as the number of new cases of COVID-19 divided by an estimate of the number of school-aged children residing in Davidson County for three referent study periods, reflecting date ranges of in-person school for School A (early and late) and School B. Two referent study periods for School A were considered, one beginning on September 9 (early; when the first students returned to in-person school) and a second beginning on October 12, 2020 (late; final date at which all students returned to in-person school). Both study periods were considered through February 12, 2021.

The referent study period for School B was August 18, 2020 through February 12, 2021. We additionally evaluated whether the number of weekly COVID-19 cases among students was related to weekly rates of COVID-19 among school-aged children in Davidson County using Pearson correlation coefficients.

The attack rate of COVID-19 among students quarantined due to in-school exposure to an infectious case and number of missed in-person school days for quarantined students were additionally estimated. The number of missed in-person school days per student was estimated based on the start date of their quarantine, considering the school protocol defined quarantine length at that time and subtracting out weekend days. Ninety-five percent confidence intervals (CI) for all cumulative incidence and attack rate calculations were estimated using the exact method.(13) A two-sided p-value less than 0.05 was used to indicate statistical significance.

Analyses were carried out using R (version 4.0.2; R Foundation for Statistical Computing, Vienna, Austria).

## RESULTS

From the start of the COVID-19 pandemic through February 12, 2021, there were 78,284 cases of COVID-19 in Davidson County, including 6,926 (8.8%) among children aged 5-18 years. The number of new cases and cumulative incidence of COVID-19 for each of the three considered study periods (School A early, School A late, School B) are reported in Table 1. The number of new cases of COVID-19 per week in schools was not strongly correlated to weekly case rates in Davidson County (r=0.37, p=0.06) (Figure 1; Figure 2).

**Table 1.** Number of cases of COVID-19 and cumulative incidence of COVID-19 at School A and School B from August 18, 2020 through February 12, 2021, and three reference periods of COVID-19 cases and cumulative incidence among school-aged children.

| **<u>School data</u>** |  |  |  |  |  |  |  |  |  |
| --- | --- | --- | --- | --- | --- | --- | --- | --- | --- |
|  | **N** | **Number of cases** |  |  |  | **Cumulative incidence (95% CI)** |  |  |  |
|  |  | School | Home | Community | Total | School | Home | Community | Total |
| **School A** |  |  |  |  |  |  |  |  |  |
| Lower | 345 | 0 | 9 | 2 | 11 | 0 (0.0, 1.1) | 2.6 (1.2, 4.9) | 0.6 (0.1, 2.1) | 3.2 (1.6, 5.6) |
| Middle | 299 | 4 | 14 | 5 | 23 | 1.3 (0.4, 3.4) | 4.7 (2.6, 7.7) | 1.7 (0.5, 3.9) | 7.7 (4.9, 11.3) |
| High | 395 | 7 | 14 | 16 | 37 | 1.8 (0.7, 3.6) | 3.5 (2, 5.9) | 4.1 (2.3, 6.5) | 9.4 (6.7, 12.7) |
| Total | 1039 | 11 | 37 | 23 | 71 | 1.1 (0.5, 1.9) | 3.6 (2.5, 4.9) | 2.2 (1.4, 3.0) | 6.8 (5.4, 8.7) |
| **School B** |  |  |  |  |  |  |  |  |  |
| Lower | 209 | 0 | 11 | 3 | 14 | 0 (0.0, 1.7) | 5.3 (2.7, 9.2) | 1.4 (0.3, 4.1) | 6.7 (3.7, 11) |
| Middle | 153 | 0 | 9 | 4 | 13 | 0 (0.0, 2.4) | 5.9 (2.7, 10.9) | 2.6 (0.7, 6.6) | 8.5 (4.6, 14.1) |
| Total | 362 | 0 | 20 | 7 | 27 | 0 (0.0, 1.0) | 5.5 (3.4, 8.4) | 1.9 (0.8, 3.9) | 7.5 (5.0, 10.7) |
| **Total** | 1401 | 11 | 57 | 30 | 98 | 0.8 (0.4, 1.4) | 4.1 (3.1, 5.2) | 2.1 (1.4, 3.0) | 7.0 (5.7, 8.5) |

| **<u>Davidson county reference data</u>** |  |  |  |
| --- | --- | --- | --- |
| **Reference period** | **N** | **Number of cases** | **Cumulative incidence (95% CI)** |
| School A<sub>early</sub> (September 9, 2020 through February 12, 2021) | 111,758 | 4695 | 4.2 (4.1-4.3) |
| School A<sub>late</sub> (October 12, 2020 through February 12, 2021) | 111,758 | 4262 | 3.8 (3.7-3.9) |
| School B (August 18, 2020 through February 12, 2021) | 111,758 | 4920 | 4.4 (4.3-4.5) |

**Abbreviations:** CI: confidence interval
**Footnote:** Davidson county reference periods were calculated for three time periods: School A<sub>early</sub> is the period starting September 9, 2020 through February 12, 2021, corresponding to the first date that any students attended school in-person at School A. School A<sub>late</sub> is the period starting October 12, 2020 through February 12, 2021, corresponding to the date the last group of students returned to in-person school at School A. The School B reference period is from August 18, 2020 through February 12, 2021, given all students were attending in-person school during that time.

**Figure 1.**
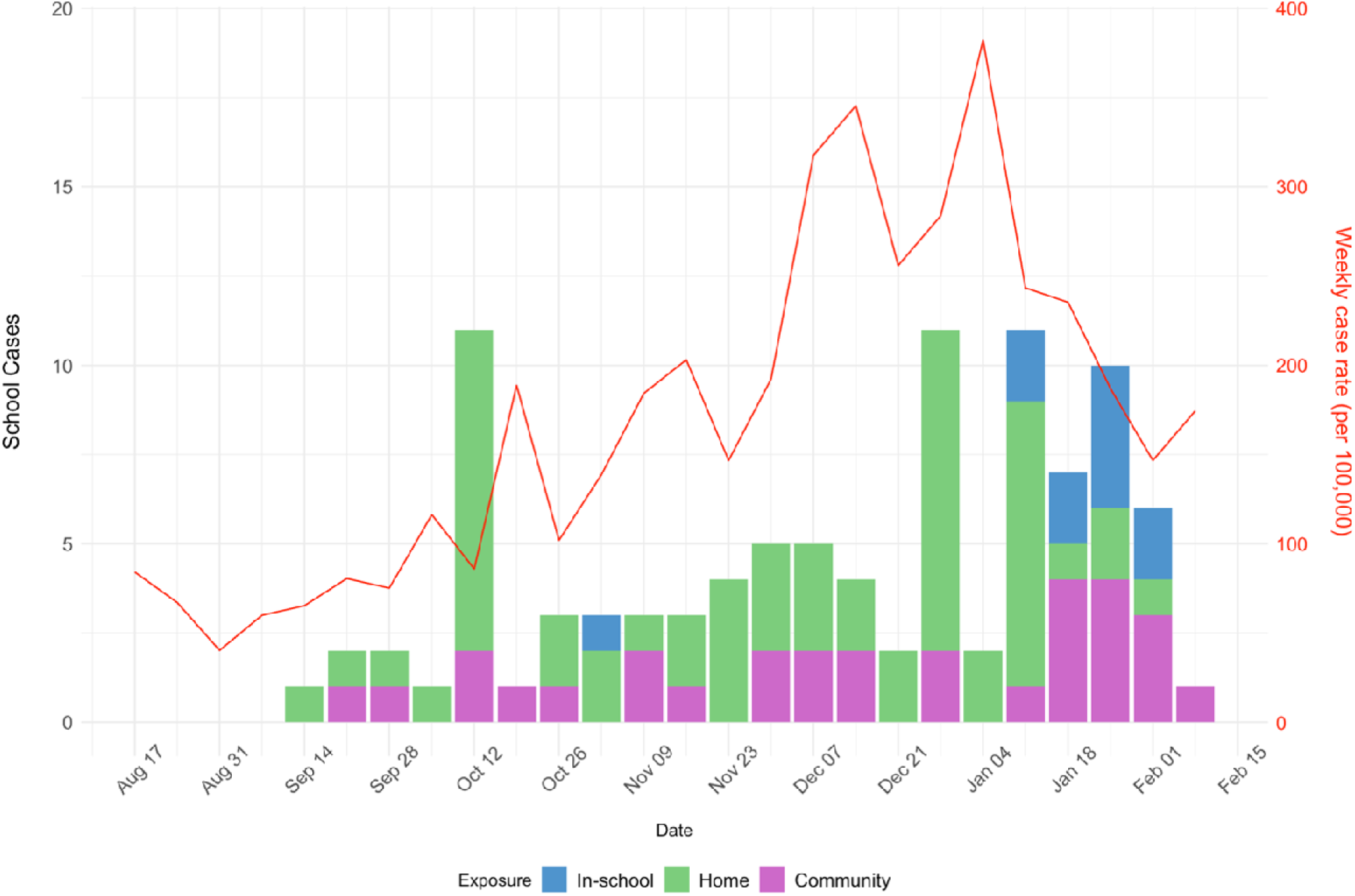
Number of cases of COVID-19 at School A and School B (left y-axis) by week (x-axis) and exposure setting (color), compared to weekly case rate of COVID-19 among school-aged children in Davidson County (right y-axis) from August 18, 2020 through February 12, 2021.

**Figure 2.**
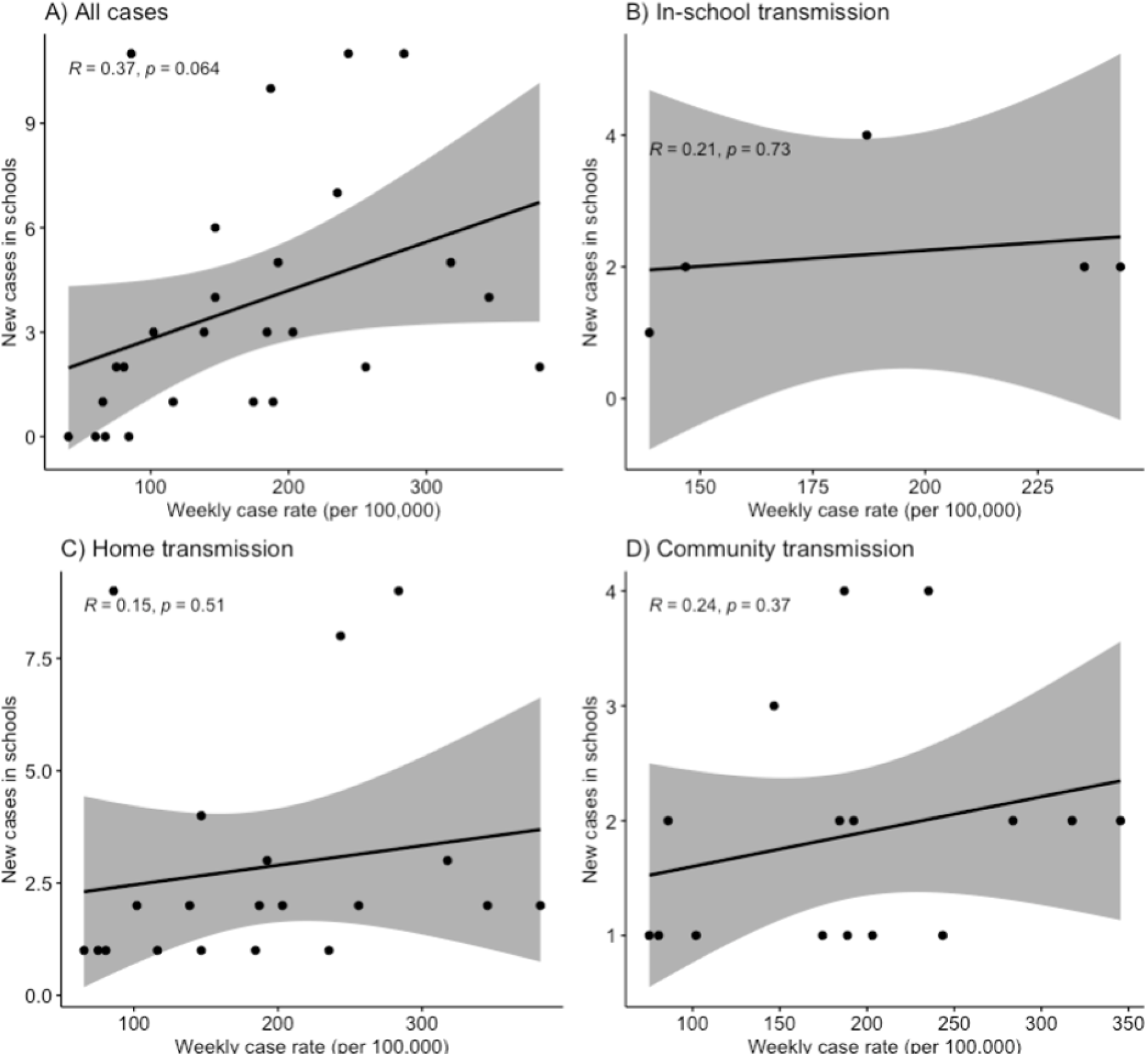
Correlation plots of new cases in schools per week compared to weekly case rate (per 100,000 school-aged children) in Davidson County for A) all cases, B) cases acquired in-school, C), cases acquired at home, and D) cases acquired in the community.

During the study period, there were 98 cases of COVID-19 among 1,401 students attending school in-person at School A and School B, corresponding to an overall cumulative incidence of 7.0% (95% CI: 5.7-8.5) [cumulative incidence was 6.8% (95% CI: 5.4-8.7) and 7.5% (95% CI: 5.0-10.7) for School A and B, respectively]. Overall, 25 cases (26%) were in lower school, 36 (37%) in middle school, and 37 (38%) in high school, corresponding to a cumulative incidence in each grade group of 4.5% (95% CI: 2.9-6.6), 8.0% (95% CI: 5.6-10.9), and 9.4% (95% CI: 6.7-12.7), respectively (Table 1). The majority (85%) of students were symptomatic at some point during their illness, though the rate of symptomatic illness was lower among younger students (p<0.01) (Table 2). Over half of cases were linked to household transmission (n=57, 58%), followed by community transmission (n=30, 31%), and few cases were linked to in-school transmission (n=11, 11%). Community transmission was more common among older age groups, whereas younger age groups were primarily infected at home (p<0.01). Of the 11 students who were infected in-school, the index case was a teacher in two instances (18%) and another student in nine instances (82%).

**Table 2.**
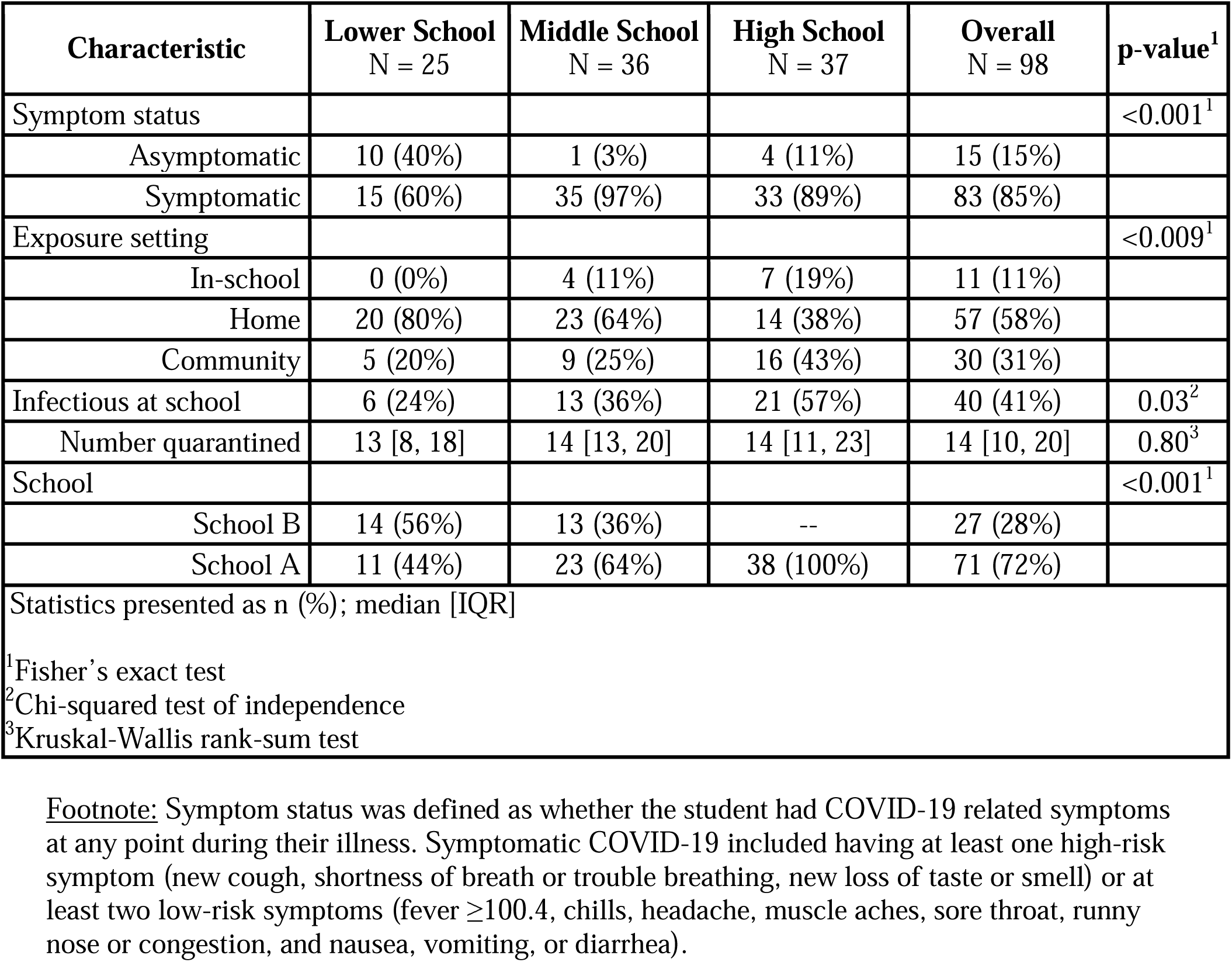
Characteristics of COVID-19 cases by school division.

Forty students were proximately known to be at school during their infectious period, resulting in quarantine of 619 other students who were deemed to be close contacts, corresponding to more than 5000 person-days of missed school. The median number of students quarantined from a single infectious case was 14 (interquartile range (IQR): 10-20). (Table 2) Among the quarantined students, only five tested positive for SARS-CoV-2 during their quarantine, corresponding to a very low primary attack rate of 0.8% (95% CI: 0.3-1.9). None of these students had other suspected exposure to COVID-19 near this time. The five cases all occurred within 7 days of exposure at school (range: 2-7).

Overall, there were 14 case clusters (two epidemiologically linked cases who were diagnosed within 14 days of each other) involving students and employees at School A and School B (Figure 3). Of those, 6 involved in-school transmission: one with transmission from staff to staff, one with transmission from staff to student, and four with transmission from one student to another student. There were no identified instances of student-to-staff transmission and no transmission linked to participation on school sports teams, which were only taking place at School A.

**Figure 3.**
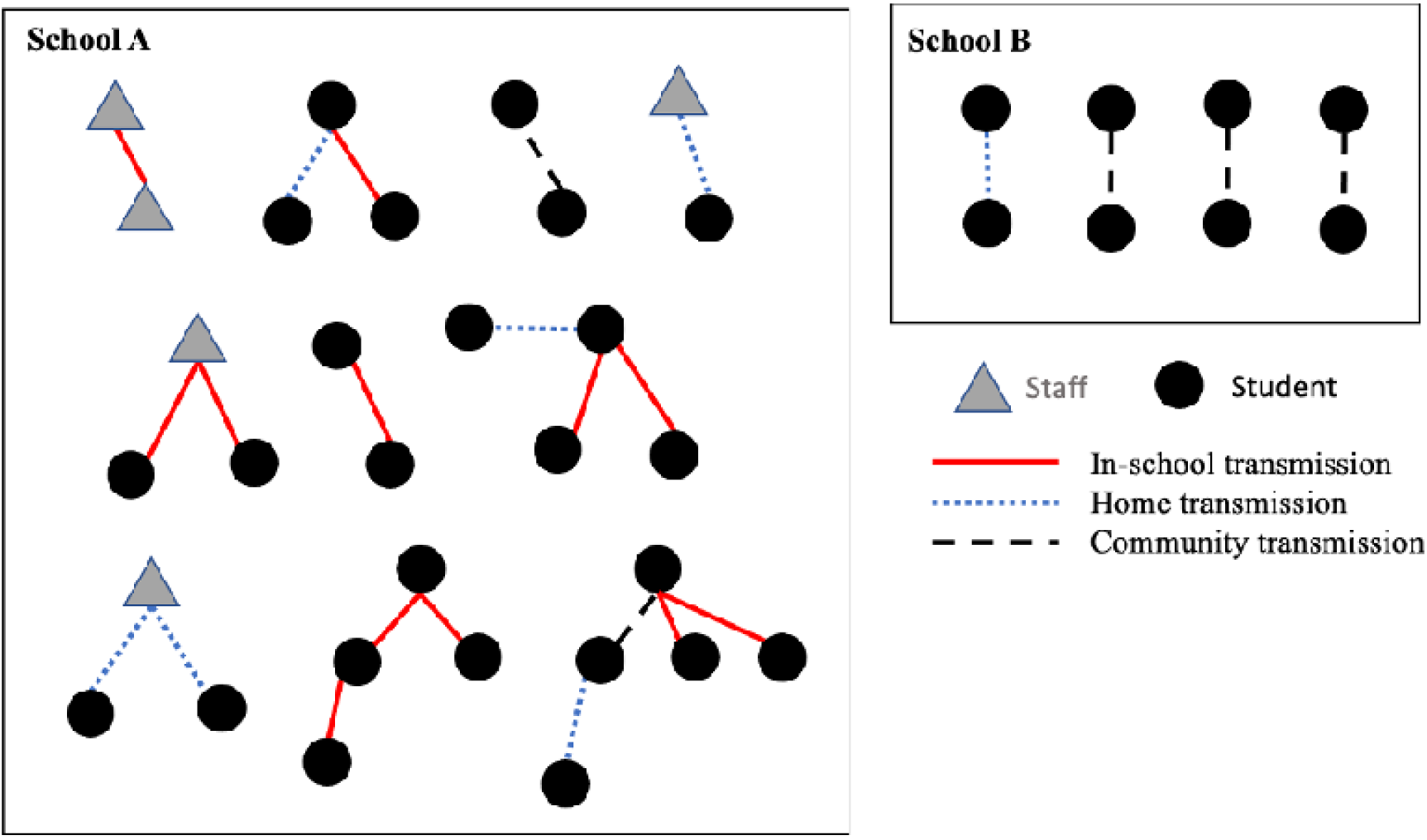
Diagram of case clusters among students and staff at School A and School B, including transmission route.

## DISCUSSION

These findings demonstrate that most COVID-19 cases among school-aged children at two independent schools in Nashville, TN with strict mitigation measures and quarantine protocols were not due to in-school transmission. These results reinforce earlier studies suggesting that student-to-teacher transmission is rare and, overall, in-school transmission is limited with adherence to strict mitigation protocols.(1,14) Specifically, a recent large study found that in-person schooling was associated increased COVID-19 rates among household members of those attending school in-person, but that the risk was greatly reduced and no longer significant when seven or more mitigation measures were used in schools.(15) The most important mitigation measures to reduce risk were daily symptom screening and mask mandates, which were employed at both schools in this study. We provide detailed reporting of the mitigation protocols used at each school, which may be helpful to other schools seeking to decrease transmission risk. Alternatively, unlike a previous report, our results do not support a strong association between cases reported to school settings and local transmission rates, but analyses may have been limited by sample size.(5,16,17)

We further highlight that many students were quarantined, corresponding to an exceedingly high number of missed in-person school days during the study period, despite minimal in-school transmission risk when multiple mitigation measures are used; the primary attack rate among quarantined students was less than 1%. Generally, there exists limited data on the appropriate strategy for quarantining and how to define close contacts aside from Centers for Disease Control and Prevention (CDC) guidelines. Though the updated operational strategy for elementary, middle, and high schools now allows for a minimum of 3 feet distancing in classrooms when universal mask use is in place, it does not specify any changes to the definition of close contact or advise on quarantine protocols.(18) A recent report from several schools in Utah found that a modified quarantine protocol, whereby school contacts were not quarantined despite meeting the CDC definition of close contact when universal mask use was in effect, was not associated with additional in-school transmission.(19) These investigators also estimated that it saved over 1,200 student in-person learning days among the 158 students who were considered close contacts but not quarantined due to consistent mask use during their study period. Based on these data and with additional support from Lessler and colleagues on how layered mitigation measures reduce transmission risk, careful consideration is warranted regarding how stringent quarantine protocols need to be in schools when daily symptom screening, universal masking, and other mitigation measures in effect.(15) This is increasingly relevant as schools define their re-opening plans for next school year and as community rates of vaccination increase. It is additionally worth considering what impact vaccinations will have on in-school transmission, given the likelihood that many adolescents will be immunized prior to the start of the 2021-2022 school year, which may allow for additional modifications to mitigation measures and quarantine protocols.

We note that the cumulative incidence of COVID-19 at School A and School B during the study period was slightly higher than reported in children in Davidson County, which can likely be explained by substantial differences in data capture via passive surveillance (Davidson County data) and active surveillance (school data). It has been widely reported that population-based COVID-19 surveillance underestimates SARS-CoV-2 infection rates.(20,21) In this study, students were encouraged to seek testing following known/suspected exposure or when they exhibited COVID-like symptoms, which likely led to improved capture of SARS-CoV-2 infection, though still less than that of universal asymptomatic serologic testing. Davidson County public schools were operating remotely during the study period, which may have resulted in less frequent COVID-19 testing due to less perceived risk and fewer social interactions. Additionally, it is possible that the students and families who comprise the study population represent a demographic that is more likely to access COVID-19 testing than the general population. Similar findings were recently reported by Zimmerman and colleagues, who found a higher number of COVID-19 clusters among students attending private schools, compared to those in public schools.(14)

### Limitations

There are several limitations of this study to consider. First, the data were retrospectively reviewed for this report. Although detailed data were collected at each school throughout the school year, the final data were not adjudicated until the time of this analysis. Some cases may have gone undiagnosed or unreported during the study period, particularly if they were asymptomatic or only mildly symptomatic and not tested for SARS-CoV-2, which is more common among younger children.(1) Second, data from Davidson County, which were used as a comparator group may not reflect the current distribution of school-aged children in Tennessee, given their reliance on 2019 census data, and may underestimate the rate of COVID-19 among school-aged children given their reliance on passive surveillance. Surveillance data commonly underreports disease activity, given reliance on passive data collection, which may explain why we see a slightly higher rate of COVID-19 reported to the two independent schools, where active case finding occurred. Finally, data for this study were collected through early February 2021, prior to widespread rollout of COVID-19 vaccinations and when there were limited local data on variants of concern. It is not yet clear what impact household and teacher vaccinations will have on COVID-19 transmission among children or in schools, but we expect that it will have beneficial effects on reducing in-school transmission, especially if mitigation measures remain in place. Conversely, if disease variants that are more transmissible become the dominant strain in the U.S., additional caution and mitigation measures may need to be considered.

### Conclusions

In conclusion, these findings add to the growing literature that COVID-19 transmission in schools is limited when strict mitigation measures are used, despite high levels of community transmission at times. Low rates of transmission in our study and the low primary attack rate during quarantine suggest that quarantining of students following in-school exposure to SARS-CoV-2 infection may be unnecessary when universal masking and other strict mitigation measures are in place.

## Data Availability

All data produced in the present study are available upon reasonable request to the authors

## IMPLICATIONS FOR SCHOOL HEALTH

Results of our study indicate that in-person schooling is safe during the COVID-19 pandemic when universal masking and other strict mitigation measures are in place, even during periods of high community transmission. Emphasis should be placed on adherence to universal masking in the school setting, particularly for unvaccinated students and employees. With strict mask adherence, schools could consider modified or no quarantine for students exposed to COVID-19, in accordance with current CDC guidelines, which would decrease missed school days (and learning opportunities) for students and missed work days for their families.

### Human subjects approval statement

This study was approved by the Vanderbilt University Medical Center Institutional Review Board.

### Funding support

This work was supported by the Dolly Parton COVID-19 Research Fund to Vanderbilt University Medical Center; the Centers for Disease Control and Prevention Leadership in Epidemiology, Antimicrobial Stewardship and Public Health (LEAP) fellowship, sponsored by the Society for Healthcare Epidemiology of America (SHEA), Infectious Diseases Society of America (IDSA) and Pediatric Infectious Diseases Society (PIDS) to S.K. This work was additionally supported by the National Institutes of Allergy and Infectious Diseases [F31AI152614-01A1 to L.S.P]. Its contents are solely the responsibility of the authors and do not necessarily represent the official views of the National Institutes of Health. The other authors received no external funding for this study.

## Acknowledgements

We would like to thank the school directors, health teams, medical advisory boards and families, without whom this project would not have been possible

## Contributions

LSP, PFR, SAM, LL, and SEK conceptualized and designed the study

LSP, LL, and SEK collected data, carried out initial analysis, drafted the initial manuscript, and reviewed and revised the manuscript.

KME, RB, SAM, and DMA reviewed and revised the manuscript, contributing critical intellectual input.

All authors approved the final manuscript as submitted and agree to be accountable for all aspects of work.

