## Supplementary File 1 for "Minimal in-school SARS-CoV-2 transmission with strict mitigation protocols at two independent schools in Nashville, TN"

**Supplementary File 1. COVID-19 protocols**

**Data collection protocols**

Data was collected via phone or email interview by the school health team. At School A, the school health team is led by a medical doctor (MD) and a registered nurse (RN). At School B, the school health team was led by a registered nurse (RN). The school health team at each school tracked student attendance. Families were encouraged to contact the school nurse for guidance on testing if their child was absent due to confirmed or suspected exposure to SARS-CoV-2 infection or symptoms consistent with SARS-CoV-2 infection (defined below). If the family did not contact the school during a student’s absence, the nurse would follow-up with that family via phone or email. All COVID-19 related health reports were adjudicated by the COVID-19 advisory team members at each school, including an epidemiologist at School A and an infectious disease physician at School B, both of whom have faculty appointments at Vanderbilt Medical Center.

**Definitions**

COVID-19 symptoms: symptoms common to SARS-CoV-2 infection were grouped as “high-risk” and “low-risk”. Families were instructed to seek healthcare and COVID-19 testing if their child was showing at least one high-risk symptom or two low-risk symptoms. High risk symptoms included: new cough, shortness of breath or trouble breathing, new loss of taste or smell. Low risk symptoms included fever ≥100.4 or chills, headache, muscle aches, sore throat, runny nose or congestion, or nausea, vomiting, or diarrhea.

Confirmed COVID-19: At both schools, a confirmed COVID-19 diagnosis (SARS-CoV-2 infection) required a positive polymerase chain reaction (PCR) test. Antigen testing alone was not sufficient to be considered as a COVID-19 diagnosis.

Suspected COVID-19: Suspected COVID-19 included individuals with symptoms consistent with COVID-19, but without PCR-confirmed SARS-CoV-2 infection, either awaiting SARS-CoV-2 test result or choice to not be tested for COVID-19.

Close contact: A close contact was someone who spent at least 15 minutes within 6 feet of an individual with confirmed or suspected COVID-19.

**Mitigation strategy**

Symptom and exposure screening: At each school, students, faculty, and staff were screened daily for symptoms consistent with COVID-19 or exposure to COVID-19. The symptom screening questions for each school are listed in the table below:

| **School A** | **School B** |
| --- | --- |
| 1. Within the last 14 days, has this student been in close contact with a person with confirmed COVID-19? 2. Is this student experiencing a new cough, difficulty breathing, or shortness of breath? 3. Is this student experiencing a change from baseline for ONE or more of the following: congestion, runny nose, sore throat, headache, fatigue or muscle aches? 4. Within the last 48 hours, has this student had a fever (temperature of 100.4 or higher)? 5. Within the last 24 hours, has this student had a new loss of smell or taste? 6. Within the last 24 hours, has this student had diarrhea or vomiting? | 1. Have you or anyone in hour household been around anyone who has tested positive for COVID-19? 2. Do you or anyone in your household exhibit any of the major COVID symptoms, including a loss of sense of taste or smell, new cough or shortness of breath or trouble breathing? 3. Do you or anyone in your household feel at all sick? 4. Are you or is anyone in your household waiting for results from a COVID-19 test? |

At School A, the symptom and exposure screening was completed by a web-based application prior to attending school, which were sent directly to the school health team. If the respondent answered “Yes” to any question, they were instructed not to attend school.

At School B, the symptom and exposure screening questions were administered at student drop off. Teacher/staff had daily temperature checks and were expected to let the school nurse know if they had exposure to COVID-19 or COVID-19 related symptoms.

At both schools, school visitors were limited, and if allowed on a case by case basis, were subjected to the same symptom and exposure screening questions as the students at that school.

Universal mask use: Students, faculty, and staff at both schools always wore masks on school property except for during lunch. During lunch, students sat outside and spaced 3-6 feet apart. If the weather did not permit outdoor lunch, then the students sat spaced 3-6 feet apart at the desk and ate in silence.

Strict hand hygiene: Students, teachers, and staff were encouraged to regularly wash their hands with soap and water or to use hand sanitizer if unable to wash hands at that time. They were additionally instructed to cough or sneeze into a tissue or elbow and to clean hands after.

Physical distancing: In classrooms at both schools, the desks were spaced 3-6 feet apart. When desks were spaced fewer than 6 feet apart, plexiglass barriers were installed between desks to further reduce transmission risk.

Class cohorts: For lower school and middle school, students remained with their class cohort for all classes and there was no mixing of class cohorts during the day. For high school students, there was very little mixing of class cohorts for grades 9 and 10, and slightly more, but still minimal mixing in grades 11 and 12.

Air filtration and sanitation: School A upgraded their HVAC and air filtration system prior to the school year. The school installed bipolar ionization systems that produce positively and negatively charged particles to remove viruses, including SARS-CoV-2, circulating within campus buildings. The school also continued with traditional filter systems in classroom air handlers on a regular replacement schedule. Teachers sanitized all surfaces between classes. School B added UV lights to their existing HVAC units and changed the filters to MERV 13, which filters out particles less than one micron in size. Teachers at School B additionally had UV wands to disinfect surfaces or other materials in their classrooms.

Sports teams: At School A, sports teams practiced following guidelines set by the American Academy of Pediatrics, which included universal masking during practice and competition?, frequent handwashing/sanitation, and active symptom and exposure screening. Sports teams did not convene at School B during the study period.

**Contact tracing and quarantine**

Families were advised to keep all students at home starting from the first sign or symptom of COVID-19 in a household member or suspected contact.

The close contacts of students, teachers, or staff, who attended school during their infectious period (48 hours prior to symptom onset or positive PCR test confirming SARS-CoV-2 infection) were required to quarantine. When it was not possible discern the exact distance between students or contact patterns, whole classrooms were quarantined.

Length of quarantine: At School A, the quarantine period was 14 days from positive test or onset of symptoms through December 5, 2020, when CDC issued revised quarantine guidelines. After that, the period of quarantine was reduced to 10 days. At School B, the quarantine period was 14 days through December 5, 2020. After that, the quarantine period changed to 10 days for all close contacts, with the option to reduce to 7 days if that individual remained symptom free and had a negative SARS-CoV-2 PCR test after day 5. Any student who developed symptoms during their quarantine was encouraged to get tested for SARS-CoV-2 and not return to school until test returned negative, symptoms resolved or their isolation period ended.

**Staggered re-opening**

School A implemented a staggered re-opening plan to gauge in-school transmission and ensure safety of students, teachers, and staff. The following schedule was used:

| September 9-11 | Kindergarten gradually returned |
| --- | --- |
| September 14-18 | Grades 1-2 gradually returned |
| September 21-25 | Grades 3-8 gradually returned |
| October 1 | Grade 10 returned |
| October. 5 | Grade 11 returned |
| October 6 | Grade 9 returned |
| October 12 | Grade 12 returned |

Gradual return means some students were in-school, some were remote, starting with half-days for a portion of students.
